# Feasibility, Implementation and Early Adoption of an Omega-3 Test-and-Treat Program to Reduce Preterm Birth

**DOI:** 10.1101/2025.06.12.25329455

**Authors:** Karen P Best, Celine Northcott, Lucy A Simmonds, Philippa Middleton, Lisa N Yelland, Vanessa Moffa, Khoa Lam, Penelope Coates, Cornelia Späth, Carol Wai-Kwan Siu, Karen Glover, Rhiannon Smith, Robert Gibson, Maria Makrides

## Abstract

**Objective:** To evaluate the feasibility and early adoption of the Omega-3 Test-and-Treat Program, a targeted intervention to reduce preterm birth in women with low omega-3 levels, implemented within routine antenatal care.

**Design:** A prospective implementation study using the Quality Enhancement Research Initiative (QUERI) framework, conducted between April 19, 2021, and June 30, 2022.

**Setting:** Antenatal care settings in South Australia, leveraging the South Australia (SA) Pathology, South Australian Serum Antenatal Screening (SAMSAS) program.

**Participants:** Pregnant women with singleton pregnancies <20 weeks’ gestation undergoing antenatal screening and healthcare providers responsible for ordering and facilitating omega-3 testing.

**Intervention:** A structured program to identify women with low omega-3 levels in early pregnancy and provide evidence-based supplementation guidance to reduce the risk of preterm birth.

**Main Outcome Measures:** Program feasibility (uptake and fidelity), representativeness of early adopters compared to the broader population, adherence to program criteria (singleton pregnancies <20 weeks’ gestation), and omega-3 status distribution.

**Results:** A total of 4,801 omega-3 tests were reported by SA Pathology, with consistent uptake over time. Women tested were demographically and clinically comparable to those not tested. Among early adopters, 702 (14.7%) had low, 1,638 (34.2%) moderate, and 2,442 (51.1%) sufficient omega-3 levels. Program fidelity was high across 5057 omega-3 lab samples with 4,935 (97.6%) analysed within the standard 72-hour timeframe. Adherence to testing criteria was strong, with only 33 (0.7%) samples from pregnancies >20 weeks’ and 58 (1.2%) from multiple pregnancies.

**Conclusion:** Early evaluations show the Omega-3 Test-and-Treat Program is feasible and integrates effectively into routine antenatal care. This real-world approach demonstrates strong potential to reduce preterm birth rates through targeted nutritional intervention, supporting its scalability and broader implementation.

**SUMMARY BOX:** The known: Preterm birth is a leading cause of infant morbidity and mortality. Omega-3 supplementation reduces preterm birth risk in women with low omega-3 levels, yet no standardised protocol exists for identifying and treating omega-3 levels during pregnancy.

The new: The Omega-3 Test and Treat Program is feasible, integrates effectively into routine antenatal care, and has broad reach, with maternal characteristics of tested women consistent with those not tested.

The implications: This scalable program has the potential to reduce preterm birth rates. Strong early adoption, high fidelity, and community engagement highlight its potential for broader implementation.

## INTRODUCTION

Preterm birth, defined as delivery before 37 weeks’ gestation, is the leading cause of death among children under five in developed countries (1–3). Early preterm birth, before 34 weeks’ gestation, contributes the highest rates of mortality and morbidity (1, 2, 4). Approximately two-thirds of preterm births occur without known biological causes, underscoring the urgent need for effective prevention strategies (5, 6).

Following many years of investigation (7, 8), omega-3 long-chain polyunsaturated fatty acids (LCPUFAs) have emerged as a promising intervention to prevent preterm birth in women with low omega-3 status. Substantial evidence indicates that correcting low omega-3 levels can significantly reduce the risk of preterm birth and improve infant outcomes (9–15). Recognising this, the 2021 National Health and Medical Research Council Australian Pregnancy Care Guidelines (16) included a recommendation for omega-3 LCPUFA supplementation (800 mg docosahexaenoic acid [DHA] and 100 mg eicosapentaenoic acid [EPA] daily) for women with low omega-3 to reduce preterm birth risk (16). Similar evidence-based recommendations have since been adopted in other international guidelines (17–19).

Despite updates to clinical practice guidelines, accurately identifying women with low omega-3 levels remains challenging. Given the variability between dietary omega-3 intake and actual blood omega-3 levels, blood testing is considered the gold standard for assessing omega-3 status (20). However, a significant gap exists in current practice, with no established omega-3 testing protocol and limited expertise amongst health professionals regarding the role of omega-3 fatty acids during pregnancy.

To address this, we developed the Omega-3 Test-and-Treat Program, a world-first preventive health strategy integrating an omega-3 blood test to identify women with low omega-3 status into routine antenatal care in South Australia. The Program targets pregnant women under 20 weeks’ gestation with a singleton pregnancy, providing tailored omega-3 supplementation recommendations based on their test results. The Program’s effectiveness in reducing preterm birth will be assessed by comparing de-identified pregnancy outcomes before and after Program implementation.

This paper assesses the feasibility of embedding the Omega-3 Test-and-Treat Program into routine antenatal care by examining uptake, fidelity to program criteria, and whether early adopters are representative of the broader population undergoing early pregnancy screening. We outline pre- and early implementation activities, highlighting critical factors for integrating this strategy into routine care to reduce preterm birth.

## METHODS

This manuscript applies steps 1-4 of the Quality Enhancement Research Initiative (QUERI) framework (21) to describe the pre- and early implementation phases of the Omega-3 Test-and- Treat Program from April 19, 2021, to June 30, 2022. As the Program is ongoing, its effectiveness in reducing preterm birth is not yet reported. Reporting follows the Standards for Reporting Implementation Studies (STaRI) guidelines (22). The Program was approved by the Women’s and Children’s Health Network Human Research Ethics Committee (HREC/20/WCHN/138).

### QUERI 1: Identify and Prioritise Gaps in Care/Clinical Needs

Omega-3 LCPUFAs have emerged as a promising intervention to prevent a proportion of the 15 million preterm births that occur each year (23). A 2018 Cochrane Review involving 19,927 women, with mainly singleton pregnancies, found that routine omega-3 LCPUFA supplementation from mid-pregnancy until birth reduced the risk of early preterm birth by 42% and preterm birth by 11%. Most included trials were conducted in upper-middle or high-income countries before the contemporary widespread use of prenatal supplements containing low-dose (⁓200 mg/day) omega-3 LCPUFA (DHA and EPA) (9). Subsequent large-scale studies support these findings (10, 12, 17, 24), but indicate that omega-3 LCPUFA supplementation is most effective in women with low omega-3 status (10, 12), suggesting background intake or omega-3 status may influence preterm birth risk (10–12).

Notably, the Omega-3 to Reduce the Incidence of Prematurity (ORIP) trial randomised 5,544 women (<20 weeks’) to receive either 800 mg DHA + 100 mg EPA or control supplements (12). Secondary analyses of singleton pregnancies indicated that women with low baseline omega-3 status had higher risk of early preterm birth and were more likely to benefit from supplementation. In contrast, those with sufficient levels had a lower risk, and additional supplementation may increase their risk (13). These findings highlight the need for a precision approach to omega-3 supplementation to reduce preterm birth.

### QUERI 2: Identify Evidence-Based Practices

In 2021, Australia’s national Pregnancy Care Guidelines were updated to reflect the latest evidence (16), recommending omega-3 supplementation for women with low omega-3 status to reduce the risk of preterm birth. Similar recommendations have since been adopted by international guidelines (17–19). However, current guidelines lack a practical method for identifying women with low omega-3 status, hindering effective implementation. At the time of implementation, omega-3 testing was not routinely available in clinical practice. To address this gap, it is essential to 1) identify women with low omega-3 status early in pregnancy and 2) provide targeted evidence-based supplementation advice based on individual omega-3 levels.

These components form the foundation of the statewide Omega-3 Test-and-Treat Program, developed to support implementation of national guidelines using a precision approach to reduce preterm birth. The Program is funded through research grants and omega-3 testing has been provided at no cost to patients or referring clinicians.

### QUERI 3: Identify barriers and facilitators

To reliably identify women with low omega-3 status in early pregnancy, we selected a blood test informed by findings from the ORIP trial (12, 13), enabling identification of those most likely to benefit from supplementation. We did not consider using omega-3 dietary intake as a screening tool because of the strength of the ORIP data in the Australian context and the well-documented inconsistency between dietary omega-3 intake and blood omega-3 levels (20).

An Omega-3 Implementation Steering Committee was formed to identify barriers and develop solutions for integrating the blood test into the South Australian healthcare system. The committee comprised researchers, pathology services, local and regional health services, obstetricians, midwives, general practitioners, and consumer representatives. This collaboration revealed that leveraging an existing early pregnancy screening program, could overcome challenges in reaching health professionals and women. The South Australia (SA) Pathology, South Australian Maternal Serum Antenatal Screening (SAMSAS) program covers approximately 80% of early pregnancy screenings in the state. Integrating the Omega-3 Test-and-Treat Program into these established workflows enabled identification of women with low omega-3 status during routine first-trimester screening, ensuring timely supplementation advice for those with singleton pregnancies before 20 weeks’ gestation.

Given that previous cutoff points for identifying women with low omega-3 status were derived from whole blood (12, 13) and the SAMSAS program utilises serum, we developed and validated equations to convert the percentage of total omega-3 fatty acids between these blood fractions (25). We then established equivalent cutoff points for ‘low’, ‘moderate’, and ‘sufficient’ omega-3 status in serum, aligning with SAMSAS processes and allowing seamless integration into existing screening workflows.

### QUERI 4: Implement and Test the Intervention in Real-World Settings

#### - Embedding the Omega-3 test into routine workflows

SAMSAS referral forms were updated with an ’omega-3 status’ checkbox, enabling health professionals to easily order the test. Maternal blood samples were processed at SA Pathology’s laboratory and forwarded to the South Australian Health and Medical Research Institute (SAHMRI) Omega-3 Laboratory for analysis. Serum fatty acid concentrations were measured using gas chromatography, as previously described (25) and results were returned to SA Pathology for reporting to health professionals through established mechanisms.

Quality measures included developing standardised protocols for interlaboratory processes, covering sample receipt, omega-3 testing, result validation and reporting. Staff from both organisations underwent cross-training to support secure transfer of samples, accurate reporting, and consistent auditing.

#### - Development of tailored advice and educational materials

Health professional and consumer reference groups were established to co-design and endorse Omega-3 Test-and-Treat Program educational materials. Bi-monthly meetings gathered feedback, enabling iterative refinements to enhance information delivery. The Health Professional Group included general practitioners, obstetricians, midwives, SA Pathology staff, communications and marketing personnel and representatives from the SAHMRI Aboriginal Communities and Families Health Research Alliance. The Consumer Group comprised community members, including women with lived experience of preterm birth.

Health professionals highlighted the need for clear advice on test interpretation and supplementation, requesting alignment with other SAMSAS results to enable simultaneous delivery of advice to patients. Pathologists emphasised the importance of categorising critically low and high omega-3 levels to distinguish extreme values associated with health risk. This led to decision points for critically low (<1% of total fatty acids) and high (>10%) serum levels.

Levels below 1% are a biochemical indicator of essential fatty acid deficiency (26), while levels above 10% represent a biochemical threshold where regular omega-3 intake would be above that considered safe by the US Food and Drug Administration (27).

Consumers stressed the need for clear supplementation guidance, including what to take, when to start, and how long to continue. There was a strong preference for advice on micro-algae omega-3 sources, due to their sustainability and suitability for vegan or vegetarian women - groups over-represented among those with low omega-3 levels. Supplementation advice was based on individual omega-3 status. Women with low levels (<3.7% total serum fatty acids) were advised to commence daily supplementation with 800 mg DHA and 100 mg EPA until 37 weeks’ gestation, in line with national Pregnancy Care Guidelines. Specific brands were not endorsed, but suitable examples were available. Guidance for each omega-3 status category was available in brochures (Supplementary Material), on a user-friendly website (28), and supported through a Program hotline and email.

#### - Implementation Strategies

We employed a multifaceted implementation strategy to engage consumers and relevant healthcare providers, ensuring the consistent integration of the Omega-3 Test-and-Treat Program across diverse antenatal care settings. Educating healthcare providers was crucial but challenging due to the fragmented healthcare system, with antenatal care delivered by various independent providers across multiple sectors and regions. Extensive early outreach, supported by collaboration with key opinion leaders, health services, primary care networks, and other healthcare bodies, was essential to ensure widespread awareness and adoption. These implementation strategies, detailed in **Table 1**, leveraged multiple channels to reach a diverse audience of health professionals and consumers.

**Table 1.** Implementation Strategy Description for Health Professionals and Consumers.

| **Implementation Strategy** | **Description** |
| --- | --- |
| **For Health Professionals** |  |
| **Direct Communication** | Email from the SA Pathology Director to SAMSAS referrers about the Omega-3 Test-and-Treat Program. Email from the South Australian Chapter of the Royal Australian College of Australia and New Zealand Obstetricians and Gynaecologists to their members. |
| **Publications & Newsletters** | Published articles in state-based newsletters of professional bodies, including SA Pathology, Royal Australian College of General Practitioners, and SA Chapter of the Australian College of Midwives, to inform and engage healthcare professionals. |
| **Tailored Education Sessions** | Delivered education sessions tailored to specific audiences during hospital in-service meetings, grand rounds, and General Practitioner Obstetric Shared Care professional development sessions. Created a podcast for General Practitioners, designed to align with their continuous professional development requirements. |
| **Promotional Events** | Hosted World Prematurity Day events and related activities to highlight the importance of omega-3 testing in reducing preterm birth. |
| **Conferences & Webinars** | Delivered webinars to health professionals and presented at midwifery, General Practitioner, and pathologist conferences, targeting these key professional groups directly. |
| **For Health Professionals and Consumers** |  |
| **Educational Materials** | Distributed Health Professional and Consumer brochures to obstetric and general practitioner clinics and maternity hospitals, accompanied by updated SAMSAS referral forms featuring the omega-3 status test checkbox. |
| **Digital Resources** | Developed dedicated web pages on SAHMRI and SA Pathology websites which hosted Health Professional and Consumer information in addition to digital copies of educational materials. |
| **General Promotion & Advertising** | Ad hoc promotional interviews on various media channels, including TV, print media, radio. Promotion of the Omega-3 Test-and-Treat Program at the annual SA Pregnancy Expo. |
| **Informational Videos** | Developed and hosted informational videos tailored to specific audiences, including consumers, midwives, and medical staff. |

SA, South Australia; SAHMRI, South Australian Health and Medical Research Institute; SAMSAS, South Australian Maternal Serum Antenatal Screening

#### - Monitoring and Evaluation of Early Implementation Interventions

Regular Steering Committee meetings facilitated ongoing feedback, enabling incremental improvements to the implementation plan. Project personnel maintained detailed logs of dissemination and implementation activities. Monthly reports from SA Pathology tracked omega-3 test uptake by comparing omega-3 tests to overall SAMSAS referrals and analysing referring clinicians by location. These insights informed adjustments to improve coverage as program uptake increased. Clinical and demographic data of women were obtained from the SAMSAS referral, completed by the referring clinician. Referrals with and without omega-3 testing, were analysed, including maternal age, gestational age, weight, in vitro fertilisation (IVF) use, smoking status, ethnicity (recorded using fixed SAMSAS categories), and referral location.

Program fidelity was monitored using quality measures assessing timeliness, accuracy, and adherence to eligibility criteria. Metrics included the proportion of eligible women tested (singleton pregnancies <20 weeks’ gestation), tests reported within 72-hours, monthly audits of omega-3 status (low, moderate, sufficient), and proportion of results within expected population range (≥1% and ≤10% fatty acids). These measures supported monitoring of integration between SAHMRI and SA Pathology laboratories and adherence to the Program’s operational processes.

## RESULTS

During the initial implementation phase of the Omega-3 Test-and-Treat Program, from April 19, 2021, to June 30, 2022, omega-3 testing uptake consistently increased, resulting in a cumulative total of 4,801 tests completed, **Figure 1**. This sustained growth highlights the feasibility of integrating the Program into routine antenatal care, indicating that the process was accessible and manageable within existing healthcare workflows.

**Figure 1.**
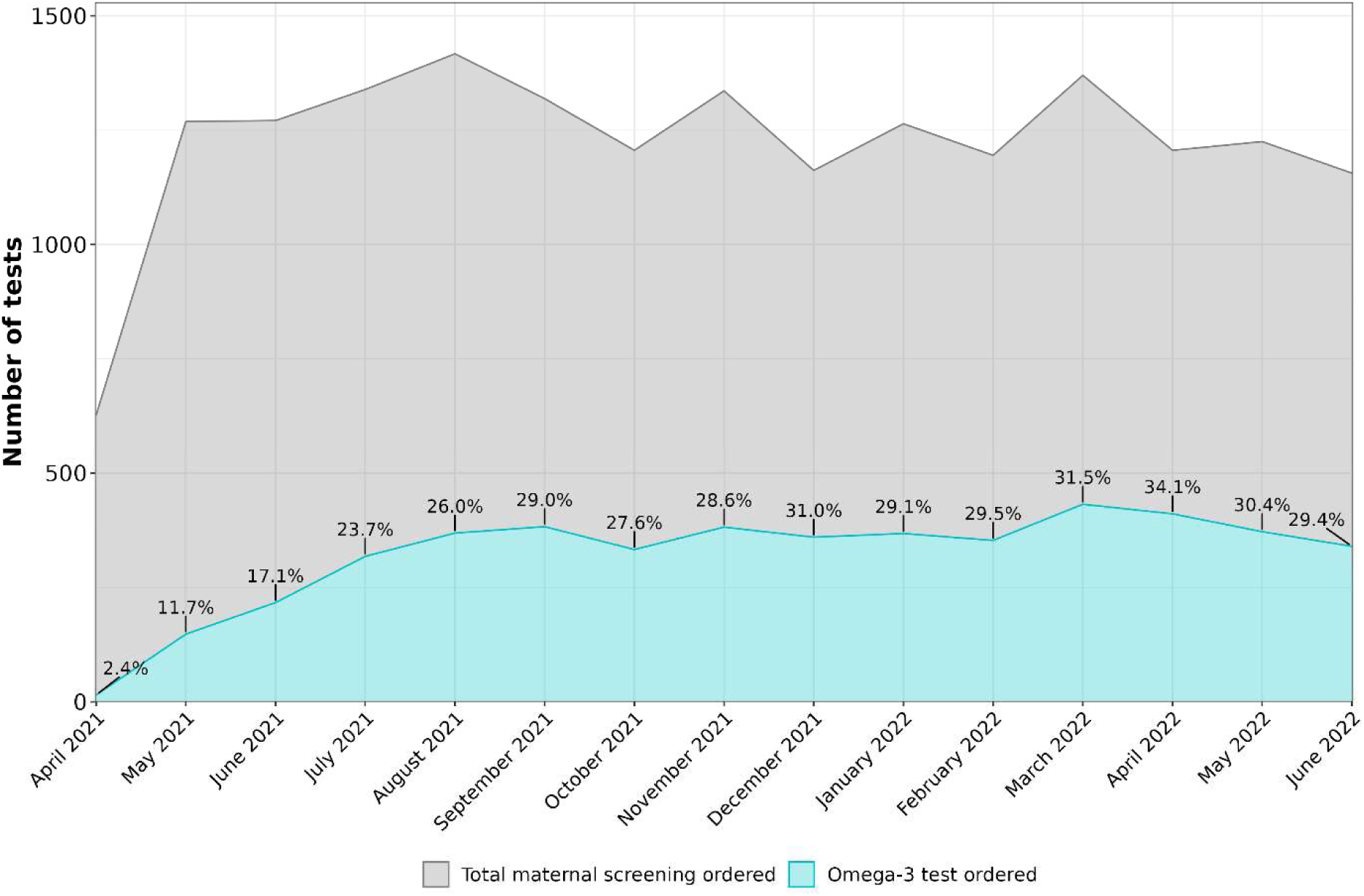
Percentage of SAMSAS referrals including omega-3 requests per month. *Note*. This figure descriptively illustrates omega-3 test uptake over time during early implementation of the Omega-3 Test-and-Treat Program in South Australia.

### Characteristics of early adopters of the Omega-3 Test-and-Treat Program

Among the 18,362 SAMSAS referral forms received by SA Pathology, 4,801 (26.1%) included an omega-3 test. Mean maternal age of women tested was 30.8 years and median gestation at testing was 12.1 weeks. Demographic and clinical characteristics were largely consistent between women who were and were not referred for omega-3 testing. Maternal age, weight, gestational age, IVF usage, and smoking rates were similar, **Table 2**.

**Table 2:** Characteristics of Early Adopters to the Omega-3 Test-and-Treat Program.

| Characteristics | Omega-3 tests<br>ordered | SAMSAS referrals<br>without omega-3 | Total Referrals<br>(with and without omega-3) |
| --- | --- | --- | --- |
| Number of tests/referrals n (%) | 4801 (26.1%) | 13,561 (73.9%) | 18,362 (100%) |
| Maternal age, mean (SD) | 30.8 (5.1%) | 30.7 (5.1%) | 30.7 (5.1%) |
| Gestational age (weeks), median (IQR) | 12.1 (11.1-13.0) | 12.4 (11.7-13.1) | 12.4 (11.6-13.1) |
| Maternal weight, median (IQR) | 69.0 (60.0-82.3) | 70.0 (60.0-83.5) | 70.0 (60.0-83.0) |
| IVF, n (%) |  |  |  |
| Yes | 255 (5.3%) | 533 (3.9%) | 788 (4.3%) |
| No | 2922 (60.9%) | 5019 (37.0%) | 7941 (43.2%) |
| Unknown | 1624 (33.8%) | 8009 (59.1%) | 9633 (52.5%) |
| Smoker, n (%) |  |  |  |
| Yes | 175 (3.6%) | 451 (3.3%) | 626 (3.4%) |
| No | 3573 (74.4%) | 6664 (49.1%) | 10237 (55.8%) |
| Unknown | 1053 (21.9%) | 6446 (47.5%) | 7499 (40.8%) |
| Ethnicity, n (%) |  |  |  |
| Caucasian<sup>1</sup> | 2659 (55.4%) | 6187 (45.6%) | 8846 (48.2%) |
| Non-Caucasian<sup>2</sup> | 745 (15.5%) | 2460 (18.1%) | 3205 (17.5%) |
| Unknown | 1397 (29.1%) | 4914 (36.2%) | 6311 (34.4%) |
| IRSAD quintiles<sup>3</sup>, n (%) |  |  |  |
| Quintile 1 – most disadvantaged | 1231 (25.6%) | 4569 (33.7%) | 5800 (31.6%) |
| Quintile 2 | 998 (20.8%) | 2667 (19.7%) | 3665 (20.0%) |
| Quintile 3 | 1070 (22.3%) | 2753 (20.3%) | 3823 (20.8%) |
| Quintile 4 | 834 (17.4%) | 1899 (14.0%) | 2733 (14.9%) |
| Quintile 5 – most advantaged | 604 (12.6%) | 1555 (11.5%) | 2159 (11.8%) |
| Unknown* | 64 (1.3%) | 118 (0.9%) | 182 (1.0%) |
| Referral location, n (%) |  |  |  |
| Metro | 3134 (65.3%) | 9836 (72.5%) | 12970 (70.6%) |
| Regional | 1604 (33.4%) | 3614 (26.6%) | 5218 (28.4%) |
| Unknown* | 63 (1.3%) | 111 (0.8%) | 174 (0.9%) |

Note: <sup>1</sup>Ethnicity categories were based on health professional selection from predefined options available on the SAMSAS form. The label “Caucasian” is used as per source form but is not endorsed terminology.
<sup>2</sup>Non-Caucasian includes all participants who did not identify as Caucasian, representing diverse racial and ethnic groups. We acknowledge the broad and simplified nature of this category.
<sup>3</sup>Ranked areas according to their relative socio-economic advantage and disadvantage based on the Socio-Economic Indexes for Areas (SEIFA), derived from clinic postcodes (42).
\*Postcode missing or not available in IRSAD datasets.
IQR, Interquartile Range; IRSAD, Index of Relative Socio-economic Advantage and Disadvantage; IVF, In Vitro Fertilisation; SD, Standard Deviation; SAMSAS, South Australian Maternal Serum Antenatal Screening

### Characteristics of early adopters by omega-3 status test result

Of the 4,801 tests, 19 had an insufficient serum sample. Of the remaining 4,782 women, 702 (14.7%) had low omega-3 levels, 1,638 (34.2%) had moderate levels, and 2,442 (51.1%) were sufficient in omega-3. Across these different omega-3 status groups, there were minimal differences in maternal age, gestational age, maternal weight, IVF status, and referral location. However, some variations were observed, such as greater representation of non-Caucasian women in the low status group (185, 26.4%) compared to the moderate and sufficient groups (226, 13.8% and 332, 13.6%, respectively). Additionally, women with low status were more likely to be smokers (51, 7.3%) compared to women with moderate (76, 4.6%) or sufficient status (47, 1.9%) **Table 3**.

**Table 3:**
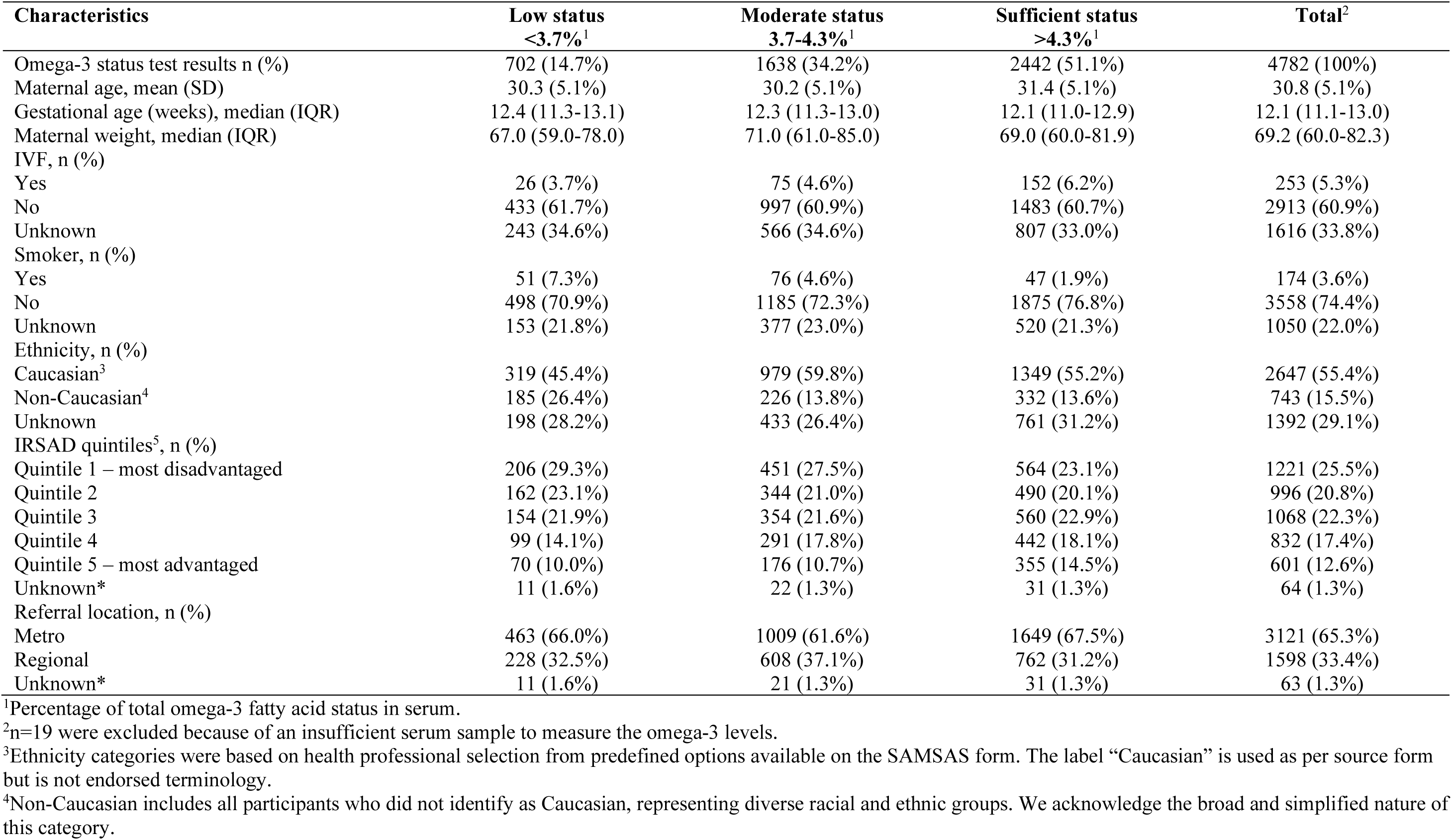
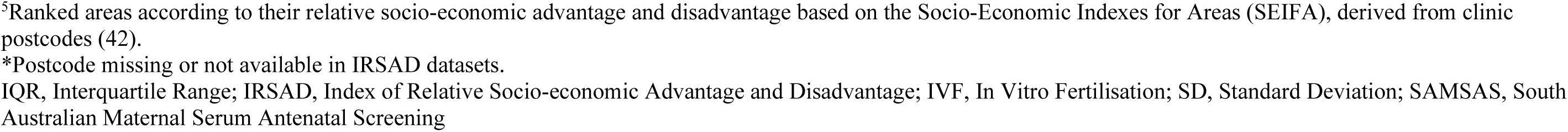
Characteristics of Early Adopters by Omega-3 Status Test Result.

| Characteristics | Low status<br><3.7%<sup>1</sup> | Moderate status<br>3.7–4.3%<sup>1</sup> | Sufficient status<br>>4.3%<sup>1</sup> | Total<sup>2</sup> |
| --- | --- | --- | --- | --- |
| Omega-3 status test results n (%) | 702 (14.7%) | 1638 (34.2%) | 2442 (51.1%) | 4782 (100%) |
| Maternal age, mean (SD) | 30.3 (5.1%) | 30.2 (5.1%) | 31.4 (5.1%) | 30.8 (5.1%) |
| Gestational age (weeks), median (IQR) | 12.4 (11.3–13.1) | 12.3 (11.3–13.0) | 12.1 (11.0–12.9) | 12.1 (11.1–13.0) |
| Maternal weight, median (IQR) | 67.0 (59.0–78.0) | 71.0 (61.0–85.0) | 69.0 (60.0–81.9) | 69.2 (60.0–82.3) |
| IVF, n (%) |  |  |  |  |
| Yes | 26 (3.7%) | 75 (4.6%) | 152 (6.2%) | 253 (5.3%) |
| No | 433 (61.7%) | 997 (60.9%) | 1483 (60.7%) | 2913 (60.9%) |
| Unknown | 243 (34.6%) | 566 (34.6%) | 807 (33.0%) | 1616 (33.8%) |
| Smoker, n (%) |  |  |  |  |
| Yes | 51 (7.3%) | 76 (4.6%) | 47 (1.9%) | 174 (3.6%) |
| No | 498 (70.9%) | 1185 (72.3%) | 1875 (76.8%) | 3558 (74.4%) |
| Unknown | 153 (21.8%) | 377 (23.0%) | 520 (21.3%) | 1050 (22.0%) |
| Ethnicity, n (%) |  |  |  |  |
| Caucasian<sup>3</sup> | 319 (45.4%) | 979 (59.8%) | 1349 (55.2%) | 2647 (55.4%) |
| Non-Caucasian<sup>4</sup> | 185 (26.4%) | 226 (13.8%) | 332 (13.6%) | 743 (15.5%) |
| Unknown | 198 (28.2%) | 433 (26.4%) | 761 (31.2%) | 1392 (29.1%) |
| IRSAD quintiles<sup>5</sup>, n (%) |  |  |  |  |
| Quintile 1 – most disadvantaged | 206 (29.3%) | 451 (27.5%) | 564 (23.1%) | 1221 (25.5%) |
| Quintile 2 | 162 (23.1%) | 344 (21.0%) | 490 (20.1%) | 996 (20.8%) |
| Quintile 3 | 154 (21.9%) | 354 (21.6%) | 560 (22.9%) | 1068 (22.3%) |
| Quintile 4 | 99 (14.1%) | 291 (17.8%) | 442 (18.1%) | 832 (17.4%) |
| Quintile 5 – most advantaged | 70 (10.0%) | 176 (10.7%) | 355 (14.5%) | 601 (12.6%) |
| Unknown* | 11 (1.6%) | 22 (1.3%) | 31 (1.3%) | 64 (1.3%) |
| Referral location, n (%) |  |  |  |  |
| Metro | 463 (66.0%) | 1009 (61.6%) | 1649 (67.5%) | 3121 (65.3%) |
| Regional | 228 (32.5%) | 608 (37.1%) | 762 (31.2%) | 1598 (33.4%) |
| Unknown* | 11 (1.6%) | 21 (1.3%) | 31 (1.3%) | 63 (1.3%) |

<sup>1</sup>Percentage of total omega-3 fatty acid status in serum.
<sup>2</sup>n=19 were excluded because of an insufficient serum sample to measure the omega-3 levels.
<sup>3</sup>Ethnicity categories were based on health professional selection from predefined options available on the SAMSAS form. The label “Caucasian” is used as per source form but is not endorsed terminology.
<sup>4</sup>Non-Caucasian includes all participants who did not identify as Caucasian, representing diverse racial and ethnic groups. We acknowledge the broad and simplified nature of this category.
<sup>5</sup>Ranked areas according to their relative socio-economic advantage and disadvantage based on the Socio-Economic Indexes for Areas (SEIFA), derived from clinic postcodes (42). \*Postcode missing or not available in IRSAD datasets. IQR, Interquartile Range; IRSAD, Index of Relative Socio-economic Advantage and Disadvantage; IVF, In Vitro Fertilisation; SD, Standard Deviation; SAMSAS, South Australian Maternal Serum Antenatal Screening

No samples recorded omega-3 levels below 1% of total fatty acids. A small number (n = 11) exceeded 10%, though only marginally and likely within the analytical margin of error.

### Measures to evaluate Program fidelity

Among the 5,057 samples received by the Omega-3 Laboratory, most were analysed and reported within the standard 72-hour timeframe (4,935, 97.6%). A small percentage of referrals were received for women beyond 20 weeks’ (33/4,801, 0.7%) or for multiple pregnancies (58/4,801, 1.2%). Together these fidelity measures indicate strong adherence to the testing criteria by health professionals.

## DISCUSSION

Early-stage evaluations demonstrate the Omega-3 Test-and-Treat Program is feasible and can be successfully integrated into routine antenatal care, offering a promising real-world strategy to reduce preterm birth rates through a targeted nutritional intervention. Central to this early success was extensive stakeholder engagement, particularly collaboration with SA Pathology, South Australia’s statewide public pathology service. By leveraging existing infrastructure and embedding omega-3 testing into existing antenatal screening pathways via the SAMSAS program, the Omega-3 Test-and-Treat Program streamlined processes and expanded its reach.

During the initial implementation period, requests for omega-3 testing steadily increased to over a quarter of all SAMSAS samples received, reflecting successful integration into existing screening protocols and strong engagement from both health professionals and women.

Further validating the Program’s broad applicability, the demographic and clinical profiles of early adopters, women tested for omega-3, closely matched those of the overall SAMSAS referral population. This consistency suggests that omega-3 testing was implemented without introducing demographic biases. The similarity between those tested and not tested for omega-3 demonstrates the Program’s extensive reach and equitable access. Omega-3 status results in this early implementation phase revealed that 702 women (14.7%) had low levels of omega-3, consistent with findings from the ORIP trial (12). This highlights a substantial portion of the population who may benefit from targeted omega-3 supplementation to reduce the risk of early preterm birth.

Analysis of women across omega-3 status groups (low, moderate, sufficient) showed minimal variations in maternal age, gestational age, maternal weight, IVF status, or referral location.

However, women who smoked were more likely to have low omega-3 status which may be attributed to the oxidative stress caused by smoking, which impacts the metabolism and levels of omega-3 fatty acids in the body (29). Women of non-Caucasian ethnicity were more likely to test low in omega-3 than Caucasian women, consistent with studies reporting ethnic differences in omega-3 fatty acid profiles (30). It is well documented that the prevalence of preterm birth is not evenly distributed, with certain populations, including Aboriginal women and socioeconomically disadvantaged populations, facing higher risks (31, 32). These findings underscore the importance of diverse input in Program design and implementation. Co-design (33) and collaboration with our Health Professional and Consumer Reference Groups, including the Aboriginal Communities and Families Health Research Alliance, helped shape the Program to meet the needs of a diverse population (34).

Program fidelity, evaluated through regular audits and feedback in line with the Consolidated Framework for Implementation Research (CFIR) (35), was crucial for maintaining consistent implementation quality. Findings indicate strong protocol adherence, with most tests conducted and reported on time and results aligning with expected population ranges. A low number of referrals outside the target population further demonstrated adherence to testing criteria. These results demonstrate the robustness of protocols and the effective collaboration between SA Pathology and SAHMRI laboratories in integrating omega-3 testing.

The early success of the Program was driven by a multifaceted approach, guided by implementation science principles, to address critical factors essential for integrating this preventive strategy into routine practice (36). Tailored education sessions, co-created educational materials and comprehensive dissemination efforts effectively raised awareness and promoted adoption (37). Accessible, well-designed materials enhanced relevance for the target audience (38), while educational meetings enhanced health professional compliance and behaviour change. Collaboration with local key opinion leaders further boosted adoption through information dissemination and endorsements.

Despite successful early adoption, limitations must be addressed. Implementing antenatal guidelines in primary care faces significant challenges, including variability in practitioner adherence, resource limitations, and the need for ongoing training and support to achieve maximal adoption (39). A key factor influencing early adoption, which was not explicitly addressed, is whether health professionals offer the omega-3 test to their patients, as they themselves could be considered early adopters of this intervention. Studies have highlighted that primary care providers often struggle with the integration of new guidelines due to time constraints, inconsistent access to up-to-date resources, and the complexity of coordinating care across different health services (39). Addressing these barriers will require sustained engagement, continuous monitoring, and iterative refinement of implementation strategies to further boost uptake. While test uptake appeared broad, the de-identified dataset used for this analysis did not include clinician-level identifiers, limiting assessment of individual ordering patterns. Missing data for some clinician-reported variables (e.g., IVF) may limit the reliability of subgroup comparisons. Encouragingly, uptake of omega-3 testing has continued to increase since the study period, indicating growing integration into routine care.

## Conclusion

Our pioneering Omega-3 Test-and-Treat Program offers a promising approach to reducing preterm birth rates through omega-3 testing and targeted supplementation. Early implementation evaluation demonstrates feasibility, with strong uptake and high levels of community engagement, supporting its potential for broader, system-level adoption. These experiences provide valuable insights to guide equitable adoption in other jurisdictions, inform future scale- up strategies and sustainable funding models, and contribute to national efforts to reduce preterm birth in Australia.

## Data Availability

The data for this study will not be shared, as we do not have permission from the participants or ethics approval to do so.

## Notes

### Competing Interest Statement

MM served as President of the International Society for the Study of Fatty Acids and Lipids (ISSFAL) from 2021 to 2024 (unpaid role). RG holds a patent titled "Stabilising and analysing fatty acids in a biological sample stored on solid media" (Patent ID AU2013209278). KB served as a member of a Preterm Birth Prevention Trial Data Safety and Monitoring Board (unpaid role). LY received funding from Societe des Produits Nestle for a separate analysis of data from the ORIP trial to identify women likely to benefit from omega-3 supplementation, which was unrelated to this work. All other authors declare no relevant disclosures.

### Funding Statement

This work was supported by a project grant from the Thyne Reid Foundation and the Hospital Research Foundation (THRF), as well as an Australian National Health and Medical Research Council (NHMRC) Centre of Research Excellence Grant (APP1135155). The ORIP trial was funded through an NHMRC project grant (APP1050468). MM and PM were supported by Australian NHMRC Investigator Grants (APP2016756 and APP1172870). KB received support from a Women's and Children's Hospital Foundation MS McLeod Postdoctoral Fellowship. The funders had no role in the design, conduct, or analysis of this work.

### Author Declarations

This work was approved by the Women's and Children's Health Network Human Research Ethics Committee (HREC/20/WCHN/138).

## REFERENCES

1. D’Apremont I, Marshall G, Musalem C, Mariani G, Musante G, Bancalari A, et al. Trends in perinatal practices and neonatal outcomes of very low birth weight infants during a 16-year period at NEOCOSUR Centers. J Pediat. 2020;225:44–50.e1.

2. Ward RM, Beachy JC. Neonatal complications following preterm birth. BJOG. 2003;110 Suppl 20:8–16.

3. World Health Organization. Born too soon: decade of action on preterm birth. Geneva; 2023.

4. Chawanpaiboon S, Vogel JP, Moller AB, Lumbiganon P, Petzold M, Hogan D, et al. Global, regional, and national estimates of levels of preterm birth in 2014: a systematic review and modelling analysis. Lancet Glob Health. 2019;7:e37–e46.

5. Ferrero DM, Larson J, Jacobsson B, Di Renzo GC, Norman JE, Martin Jr JN, et al. Cross-country individual participant analysis of 4.1 million singleton births in 5 countries with very high human development index confirms known associations but provides no biologic explanation for 2/3 of all preterm births. PloS one. 2016;11:e0162506.

6. Goodfellow L, Care A, Alfirevic Z. Controversies in the prevention of spontaneous preterm birth in asymptomatic women: an evidence summary and expert opinion. BJOG. 2021;128:177–94.

7. Olsen SF, Sørensen TA, Secher NJ, Hansen HS, Jensen B, Sommer S, et al. Intake of marine fat, rich in (n-3)-polyunsaturated fatty acids, may increase birthweight by prolonging gestation. Lancet. 1986;328:367–9.

8. Olsen SF, Sørensen JD, Secher NJ, Hedegaard M, Henriksen TB, Hansen HS, et al. Randomised controlled trial of effect of fish-oil supplementation on pregnancy duration. Lancet. 1992;339:1003–7.

9. Middleton P, Gomersall JC, Gould JF, Shepherd E, Olsen SF, Makrides M. Omega-3 fatty acid addition during pregnancy. Cochrane Database Syst Rev. 2018;11.

10. Carlson SE, Gajewski BJ, Valentine CJ, Kerling EH, Weiner CP, Cackovic M, et al. Higher dose docosahexaenoic acid supplementation during pregnancy and early preterm birth: a randomised, double-blind, adaptive-design superiority trial. EClinicalMedicine. 2021;36:100905.

11. Olsen SF, Halldorsson TI, Thorne-Lyman AL, Strom M, Gortz S, Granstrom C, et al. Plasma concentrations of long chain n-3 fatty acids in early and mid-pregnancy and risk of early preterm birth. EBioMedicine. 2018;35:325–33.

12. Makrides M, Best K, Yelland L, McPhee A, Zhou S, Quinlivan J, et al. A randomized trial of prenatal n-3 fatty acid supplementation and preterm delivery. N Engl J Med. 2019;381:1035–45.

13. Simmonds LA, Sullivan TR, Skubisz M, Middleton PF, Best KP, Yelland LN, et al. Omega- 3 fatty acid supplementation in pregnancy-baseline omega-3 status and early preterm birth: exploratory analysis of a randomised controlled trial. BJOG. 2020;127:975–81.

14. Klebanoff MA, Harper M, Lai Y, Thorp J, Jr., Sorokin Y, Varner MW, et al. Fish consumption, erythrocyte fatty acids, and preterm birth. Obstet Gynecol. 2011;117:1071–7.

15. Harper M, Thom E, Klebanoff MA, Thorp J, Jr., Sorokin Y, Varner MW, et al. Omega-3 fatty acid supplementation to prevent recurrent preterm birth: a randomized controlled trial. Obstet Gynecol. 2010;115:234–42.

16. Australian Government Department of Health: Clinical practice guidelines: pregnancy care. Canberra: Australian Government; 2021.

17. Best KP, Gibson RA, Makrides M. ISSFAL Statement number 7 - Omega-3 fatty acids during pregnancy to reduce preterm birth. Prostaglandins Leukot Essent Fatty Acids. 2022;186:102495.

18. Cetin I, Carlson SE, Burden C, da Fonseca EB, di Renzo GC, Hadjipanayis A, et al. Omega- 3 fatty acid supply in pregnancy for risk reduction of preterm and early preterm birth. Am J Obstet Gynecol MFM. 2024;6:101251.

19. Savona-Ventura C, Mahmood T, Mukhopadhyay S, Louwen F. Omega-3 fatty acid supply in pregnancy for risk reduction of preterm and early preterm birth: a position statement by the European Board and College of Obstetrics and Gynaecology (EBCOG). Eur J Obstet Gynecol Reprod Biol. 2024;295:124–5.

20. de Groot RHM, Meyer BJ. ISSFAL Official statement number 6: the importance of measuring blood omega-3 long chain polyunsaturated fatty acid levels in research. Prostaglandins Leukot Essent Fatty Acids. 2020;157:102029.

21. Stetler CB, Mittman BS, Francis J. Overview of the VA quality enhancement research initiative (QUERI) and QUERI theme articles: QUERI series. Implement Sci. 2008;3:8.

22. Pinnock H, Barwick M, Carpenter CR, Eldridge S, Grandes G, Griffiths CJ, et al. Standards for reporting implementation studies (StaRI) statement. BMJ. 2017;356:i6795.

23. Patel RM, Kandefer S, Walsh MC, Bell EF, Carlo WA, Laptook AR, et al. Causes and timing of death in extremely premature infants from 2000 through 2011. N Engl J Med. 2015;372:331–40.

24. Olsen SF, Halldorsson TI, Li M, Strom M, Mao Y, Che Y, et al. Examining the effect of fish oil supplementation in Chinese pregnant women on gestation duration and risk of preterm delivery. J Nutr. 2019;149:1942–51.

25. Simmonds LA, Yelland LN, Best KP, Liu G, Gibson RA, Makrides M. Translating n-3 polyunsaturated fatty acid status from whole blood to plasma and red blood cells during pregnancy: translating n-3 status across blood fractions in pregnancy. Prostaglandins Leukot Essent Fatty Acids. 2022;176:102367.

26. Cleland LG, James MJ, Proudman SM, Neumann MA, Gibson RA. Inhibition of human neutrophil leukotriene B4 synthesis in essential fatty acid deficiency: role of leukotriene A hydrolase. Lipids. 1994;29:151–5.

27. National Institutes of Health. Omega-3 Fatty Acids 2025 [Available from: https://ods.od.nih.gov/factsheets/Omega3FattyAcids-HealthProfessional/#h39

28. South Australian Health and Medical Research Institute. Omega-3 Test-and-Treat Program 2025 [Available from: https://sahmri.org.au/information-for-health-professionals

29. Simon JA, Fong J, Bemert Jr JT, Browner WS. Relation of smoking and alcohol consumption to serum fatty acids. Am J Epidemiol. 1996;144:325–34.

30. Reigada LC, Storch B, Alku D, Hazeltine DB, Heppelmann PG, Polokowski AR. Assessment of polyunsaturated fatty acids: a self-report and biomarker assessment with a racially and ethnically diverse sample of women. Prostaglandins Leukot Essent Fatty Acids. 2021;164:102214.

31. Wilson NA, Mantzioris E, Middleton PF, Muhlhausler BS. Influence of clinical characteristics on maternal DHA and other polyunsaturated fatty acid status in pregnancy: a systematic review. Prostaglandins Leukot Essent Fatty Acids. 2020;154:102063.

32. Kildea SV, Gao Y, Rolfe M, Boyle J, Tracy S, Barclay LM. Risk factors for preterm, low birthweight and small for gestational age births among Aboriginal women from remote communities in Northern Australia. Women Birth. 2017;30:398–405.

33. Dietrich T, Trischler J, Schuster L, Rundle-Thiele S. Co-designing services with vulnerable consumers. J Serv Theory Pract. 2017;27:663–88.

34. Doherty E, Kingsland M, Wiggers J, Wolfenden L, Hall A, McCrabb S, et al. The effectiveness of implementation strategies in improving preconception and antenatal preventive care: a systematic review. Implement Sci Commun. 2022;3(1):121.

35. Damschroder LJ, Aron DC, Keith RE, Kirsh SR, Alexander JA, Lowery JC. Fostering implementation of health services research findings into practice: a consolidated framework for advancing implementation science. Implement Sci. 2009;4:1–15.

36. Powell BJ, McMillen JC, Proctor EK, Carpenter CR, Griffey RT, Bunger AC, et al. A compilation of strategies for implementing clinical innovations in health and mental health. Med Care Res Rev. 2012;69:123–57.

37. Proctor EK, Powell BJ, McMillen JC. Implementation strategies: recommendations for specifying and reporting. Implement Sci. 2013;8:139.

38. Kingsland M, Doherty E, Anderson AE, Crooks K, Tully B, Tremain D, et al. A practice change intervention to improve antenatal care addressing alcohol consumption by women during pregnancy: research protocol for a randomised stepped-wedge cluster trial. Implement Sci. 2018;13:112.

39. Wang T, Tan JB, Liu XL, Zhao I. Barriers and enablers to implementing clinical practice guidelines in primary care: an overview of systematic reviews. BMJ Open. 2023;13:e062158.

40. Australian Bureau of Statistics. Socio-Economic Indexes for Areas (SEIFA): ABS; 2021 [Available from: https://www.abs.gov.au/statistics/people/people-and-communities/socio-economic-indexes-areas-seifa-australia/2021

